# Optimal COVID-19 epidemic control until vaccine deployment

**DOI:** 10.1101/2020.04.02.20049189

**Authors:** R. Djidjou-Demasse, Y. Michalakis, M. Choisy, M. T. Sofonea, S. Alizon

## Abstract

Since Dec 2019, the COVID-19 epidemic has spread over the globe creating one of the greatest pandemics ever witnessed. This epidemic wave will only begin to roll back once a critical proportion of the population is immunised, either by mounting natural immunity following infection, or by vaccination. The latter option can minimise the cost in terms of human lives but it requires to wait until a safe and efficient vaccine is developed, a period estimated to last at least 18 months. In this work, we use optimal control theory to explore the best strategy to implement while waiting for the vaccine. We seek a solution minimizing deaths and costs due to the implementation of the control strategy itself. We find that such a solution leads to an increasing level of control with a maximum reached near the 16th month of the epidemics and a steady decrease until vaccine deployment. The average containment level is approximately 50% during the 25-months period for vaccine deployment. This strategy strongly out-performs others with constant or cycling allocations of the same amount of resources to control the outbreak. This work opens new perspectives to mitigate the effects of the ongoing COVID-19 pandemics, and be used as a proof-of-concept in using mathematical modelling techniques to enlighten decision making and public health management in the early times of an outbreak.

## 1 Introduction

In early December 2019, an outbreak caused by a novel betacoronavirus was reported in Wuhan, the capital city of the Hubei Chinese province [3, 16]. It rapidly spread to other provinces in China and to the rest of the world. On March 17 2020, the World Health Organisation (WHO) officially declared COVID-19 a pandemic. This RNA virus is now known as SARS-CoV-2 [21] and it is reported to cause lower respiratory tract infections in humans with a variety of unspecific symptoms [3, 10, 26]. The virulence of the infection is relatively high with a case fatality ratio estimated to be in the order of 1% [5, 6, 23, 25], therefore making the virus a public health priority issue given the anticipated size of the epidemics due to the absence of pre-existing immunity.

Most countries reacted to the epidemics by rapidly implementing containment public health measures, also referred to as non-pharmaceutical interventions. China was the first country to react by putting the city of Wuhan on quarantine on January 23, 2020, shortly followed by the whole region. Other countries also implemented a national lock-down, such as France on March 17 or the United Kingdom on March 23.

Broadly speaking, there are two main ways to halt the spread of the epidemics. The first one is through natural immunisation: once a proportion 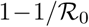 of the population has been infected (where 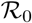 is the basic reproduction number), a ‘herd immunity’ develops and the epidemic starts to decrease [1]. Note that this herd immunity threshold is lower than the ‘worst-case scenario’, i.e. the epidemics final size if no containment measure is implemented [13]. The second option is to achieve population immunisation through vaccination. The threshold to have the epidemics decrease remains the same, but the cost in terms of mortality can be much lower. These options have been investigated by the WHO Collaborating Centre for Infectious Disease Modelling in two reports [7, 24]. The first report explores the effect of an on/off triggering of 5 types of non-pharmaceutical interventions, whereas the second report compares a containment strategy to a suppression strategy, while factoring in indirect costs of implementing the strategy. Other models have explored the efficiency of contact tracing to mitigate the epidemics [11, 12].

Here, we adopt a more modelling approach based on optimal control theory [15, 22] to determine the best strategy to implement until vaccine deployment. We therefore focus on a finite time interval [0, *T*], where *T* is the number of days required for vaccine discovery, manufacturing and deployment. Our key metric is the fraction c of decrease in 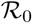 (or, equivalently, in the contact rate) obtained through non-pharmaceutical interventions, which we refer to as control intensity. Importantly, the model considers that implementing the control policy comes with a cost, which corresponds to diverting funding from important sources or to social deleterious effects, and can cause indirect mortality. Optimal control theory allows us to find the optimal control strategy over time, denoted by *c*(*t*), that minimises the cumulative sum of these costs over the [0, *T*] period of time. We compare this strategy to scenarios without any control or with more homogeneous control (i.e. with *c* alternating between constant values at a given frequency) that involve the same average effort over time as the optimal strategy. We find that the optimal control strategy outperforms the others in terms of direct and indirect mortality.

We first present the structure of our epidemiological model, then we introduce the objective function we optimise and derive the optimality condition. We then present our results and analyse them in the light of earlier models implemented in the context of pandemic influenza [14, 17] and of the ongoing COVID-19 pandemics.

## 2 The Model

### 2.1 Model overview

The model describes the epidemic dynamics of COVID-19 in a population where hosts can belong to five states: susceptible (*S*), latent *i.e*. infected but asymptomatic and not infectious (*E*), asymptomatic infectious (*A*), symptomatic infectious (*I*), recovered (*R*) and dead (*D*). Such compartments are suitable to describe the COVID-19 life cycle. Recovered hosts are assumed to be immune for life. The model is also structured by differential disease severity, since COVID-19 can cause both mild or severe infections [19, 26]. We assume that severe symptomatic infections require hospitalisation, which reduces their contagiousness. The main variables and parameters of the model are listed in Table 1 and shown in Figure 1. Next, we introduce the model and describe its parameters and general hypotheses.

**Table 1:** Model state variables and parameters

| Variables | Description |  |
| --- | --- | --- |
| $S$ | Density of susceptible individuals | |
| $E_m, E_s$ | Density of latent individuals with mild/severe infection | |
| $A_m, A_s$ | Density of asymptomatic infectious individuals with mild/severe infection | |
| $I_m, I_s$ | Density of symptomatic infectious individuals with mild/severe infection | |
| $R_m, R_s$ | Density of recovered individuals | |
| $D_{\text{COVID}}$ | Deaths directly caused by COVID-19 | |
| $D_{\text{SAT}}$ | Indirect deaths due to hospital saturation | |
| $D$ | Total deaths | |
| Parameters | Description (unit) | Value(range) [ref.] |
| $\mathcal{R}_0$ | Basic reproduction number | 2.5 (2-3) |
| $p$ | Proportion of mild infections | 0.9 (0.85-0.95) [9] |
| $\beta_I$ | Symptomatic transmission rate | Calculated<sup>a</sup> |
| $\beta_A$ | Asymptomatic transmission rate | Calculated<sup>a</sup> |
| $\xi$ | Reduced transmission factor of severe infections | 0.2 [assumed] |
| $c$ | Control effort (0,1) | |
| $\nu$ | Immigration rate ($\text{day}^{-1}$) | 2 [assumed] |
| $\varepsilon$ | Waiting rate to viral shedding ($\text{day}^{-1}$) | 1/4.2 (0.21-0.27) [16] |
| $\sigma$ | Waiting rate to symptom onset ($\text{day}^{-1}$) | 1 (0.9-1.1) [7] |
| $\gamma_m$ | Recovery rate from mild infections ($\text{day}^{-1}$) | 1/17 (0.025-0.1) [27] |
| $\gamma_s$ | Recovery rate from severe infections ($\text{day}^{-1}$) | 1/20 (0.039-0.13) [27] |
| $\mu$ | Natural mortality rate with hospital saturation ($\text{day}^{-1}$) | $10^{-5}$ [assumed] |
| $\theta$ | Case-fatality ratio in severe infections | 0.15 (0.135-0.165) [9] |
| $\alpha_{\min}$ | Lower bound disease induced mortality rate ($\text{day}^{-1}$) | Calculated<sup>b</sup> |
| $\alpha_{\max}$ | Higher bound induced mortality rate ($\text{day}^{-1}$) | $2\alpha_{\min}$ [assumed] |
| $H$ | Healthcare capacity | Variable |
| $B$ | Cost weight | Variable (0, ∞) |
| Initial conditions |  |  |
| $N_0$ | Total population | $67 \times 10^6$ |
| $I_0$ | Size of infected population | $0.01 \times H$ (variable) |

<sup>a</sup> $\beta_A$ and $\beta_I$ are calculated by (2.3)
<sup>b</sup> $\alpha_{\min}$ is calculated by (2.4)

**Figure 1:**
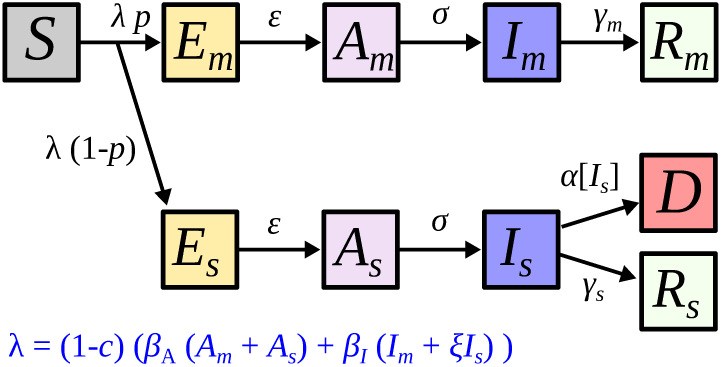
The compartmental model flow diagram. Host states are indicated by squares and transitions by arrows (along with their corresponding rates). At time *t*, the host population is composed by susceptibles (*S*). They are exposed to the virus with the force of infection *λ*. A fraction p of exposed individuals will develop mild and the remainder severe infections (subscript “*m*” and “*s*” respectively). Exposed individuals (*E*_●_) remain non-infectious during a period to viral shedding (1/*ε*). Next, they become asymptomatic infectious (*A*_●_) and will develop symptoms after a period to symptom onset (1/*σ*). Infected individuals recover from the infection at rate *γ*_●_. Only severely infected die from the infection at rate *α*[*I_s_*]. Hosts in all compartments die at a natural mortality rate *μ*[*I_s_*] that is not shown for clarity. Notations are shown in Table 1.

### 2.2 The SEAIR model with differential severity

The total population size at a given time *t* is denoted by *N* = *S+E_m_+E_s_+A_m_*+ *A_s_* + *I_m_* + *I_s_* + *R_m_* + *R_s_*, where subscript ’*s*’ and’*m*’ refer to infection severity (’severe’ or ’mild’). Without indicating the time dependency for simplicity, we define the force of infection by *λ* = (1 − c) (*β_A_*(*A_s_* + *A_m_*) + *β_I_*(*I_m_* + *ξI_s_*)), where *c* is the percentage of reduction in transmission due to public health measures at time *t*, and *β_A_* and *β_I_* are the transmission rates of symptomatic and asymptomatic infections respectively. Parameter ξ is the factor of reduction of the transmission rate of severe and symptomatic infections, which is due to being in a health care facility. A proportion *p* of exposed individuals (*λS*) move to the mild asymptomatic infection class at a rate *ε*, while the remainder (1 − *p*) move to the severe asymptomatic infection class at the same rate *ε*. We also assume that the epidemic is sustained by migration of *E_m_* and *E_s_* individuals at a constant rate *pν* and (1 − *p*)*ν* respectively. Severe and mild asymptomatic infectious individuals become symptomatic at a same rate *σ* and recover at rates *γ_s_* and *γ_m_* respectively. While mild infections are never lethal, severe cases lead to death at rate *α*[*I_s_*]. We assume that the number of severely infected hosts at a given time, *I_s_*, affects the disease induced mortality rate (if hospitals are saturated for instance). If we denote by *H* the total number of infected hosts the health care system (especially the intensive care units, ICU) can sustain, then we assume the mortality rate *α* to be such that

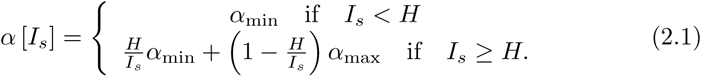

We apply the same reasoning at the whole population level by assuming that natural mortality increases because of hospital saturation. We capture this using the following function for the mortality rate *μ*:

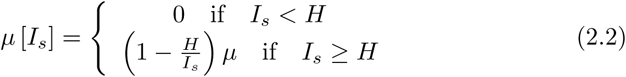

The disease-induced mortality parameter (*α*_min_) and transmission rates (*β_A_* and *β_I_*) are estimated thanks to the case-fatality ratio (θ) and the basic reproduction number 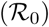 which are well known from the literature [5, 6, 16, 25]. From these and assuming *β_A_* = *β_I_*, we have:

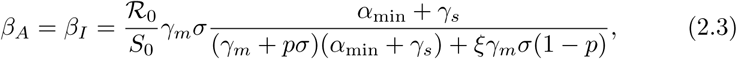

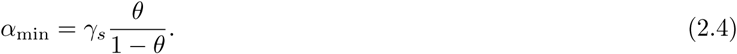

See Appendix D for details on the derivation of 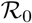.

Setting 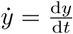, the resulting system of equations captures our model:

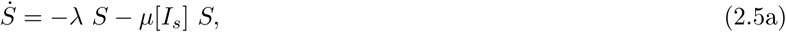

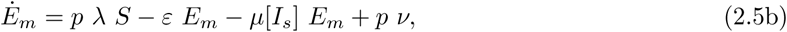

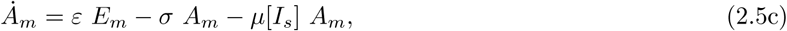

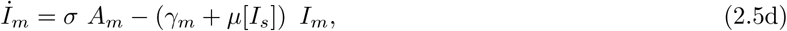

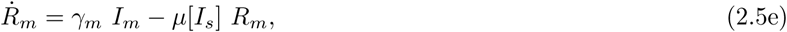

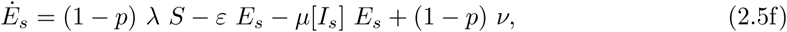

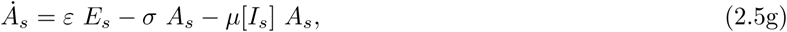

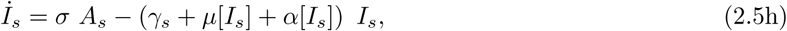

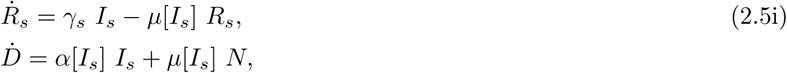

with

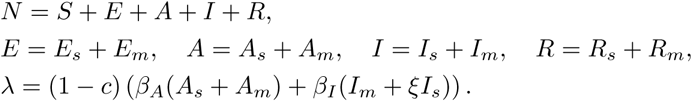

System (2.5) is coupled with the following initial conditions

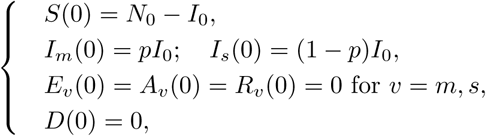

where *N*_0_ and *I*_0_ are initial total and infected populations defined in Table 1. Basic properties such as existence, positivity and boundedness solutions of (2.5) are straightforward and therefore are not detailed here.

## 3 Optimal intervention and objective function

In this section, we find the optimal control effort *c*^*^(*t*) which substantially minimizes the cumulative number of deaths until vaccine deployment. We first define the objective function to penalize and then derive the necessary optimality condition. A key step is to define the function to optimise. We assume that a successful scheme is one which reduces the number of deaths in the host population. For clarity, we separate deaths directly attributable to COVID-19 (*D*_COVID_ = *α* [*I_s_*] *I_s_*) and deaths indirectly linked to COVID-19 infection but due to the saturation of the hospital system (*D*_SAT_ = *μ* [*I*_s_] *N*).

Therefore, the control scheme is optimal if it minimizes the objective function

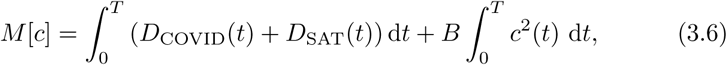

The first integral corresponds to the total number of deaths over the time period considered and the second represents the total cost associated with the implementation of the control measures. As earlier studies [14, 17], we assumed a quadratic expression in the second term associated to the control to capture non-linear costs potentially arising for intense control implementations. When the objective function is quadratic with respect to the control, differential equations arising from optimization have a known solution. Other functional forms often lead to systems of differential equations that are difficult to solve [15]. Finally, *B* is a coefficient allowing to weight the “cost” or “penalty” associated to the control implementation (*c* (*t*)). This factor is difficult to evaluate and includes many aspects ranging from economic to psychological impact. However, we can take advantage of the healthcare system capacity *H* to fix this parameter. Indeed, the greater hospital capacities, the less need there is for strict confinement (*i.e*., the control measure *c* is heavily penalized).

Our aim is to find the function *c*^*^ satisfying

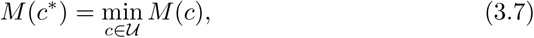

on the set 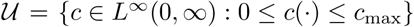, where *c*_max_ ≤ 1, and *L^∞^* the vector space of essentially bounded measurable functions.

Using Pontryagin’s maximum principle [22], the solution *c*^*^ of (3.7) is characterized by

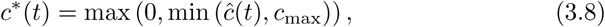

with

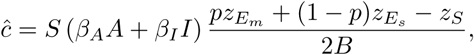

and where the adjoint functions (*z_ν_*)*_ν_∈*{*S*,*E_m_*,*E_s_*,*A_m_*,*A_s_*,*I_m_*,*I_s_*,*R_m_,R_s_*} verify (C.3).

The state system (2.5) and the adjoint system (C.3) together with the control characterization (C.4) constitute the optimality system to be solved numerically. Since the state equations have initial conditions and the adjoint equations have final time conditions, we cannot solve the optimality system directly. Instead, we use an iterative algorithm to sweep forward in time using a forward-backward sweep method [15].

## 4 Results

### Doing-nothing scenario

The model proposed here describes epidemic dynamics during several weeks in a host population before vaccine deployment. In the baseline situation, there is no public health measure to control epidemics (*c* = 0). The infection spreads rapidly and the peak is reached within few weeks with a large number of severe cases (Figure 2b). The healthcare system is quickly overwhelmed and this situation lasts for a relatively long period of time (black line in Figure 2b). Consequently, the number of deaths increases exponentially with additional deaths due to the saturation of the health system (Figure 2c,d).

**Figure 2:**
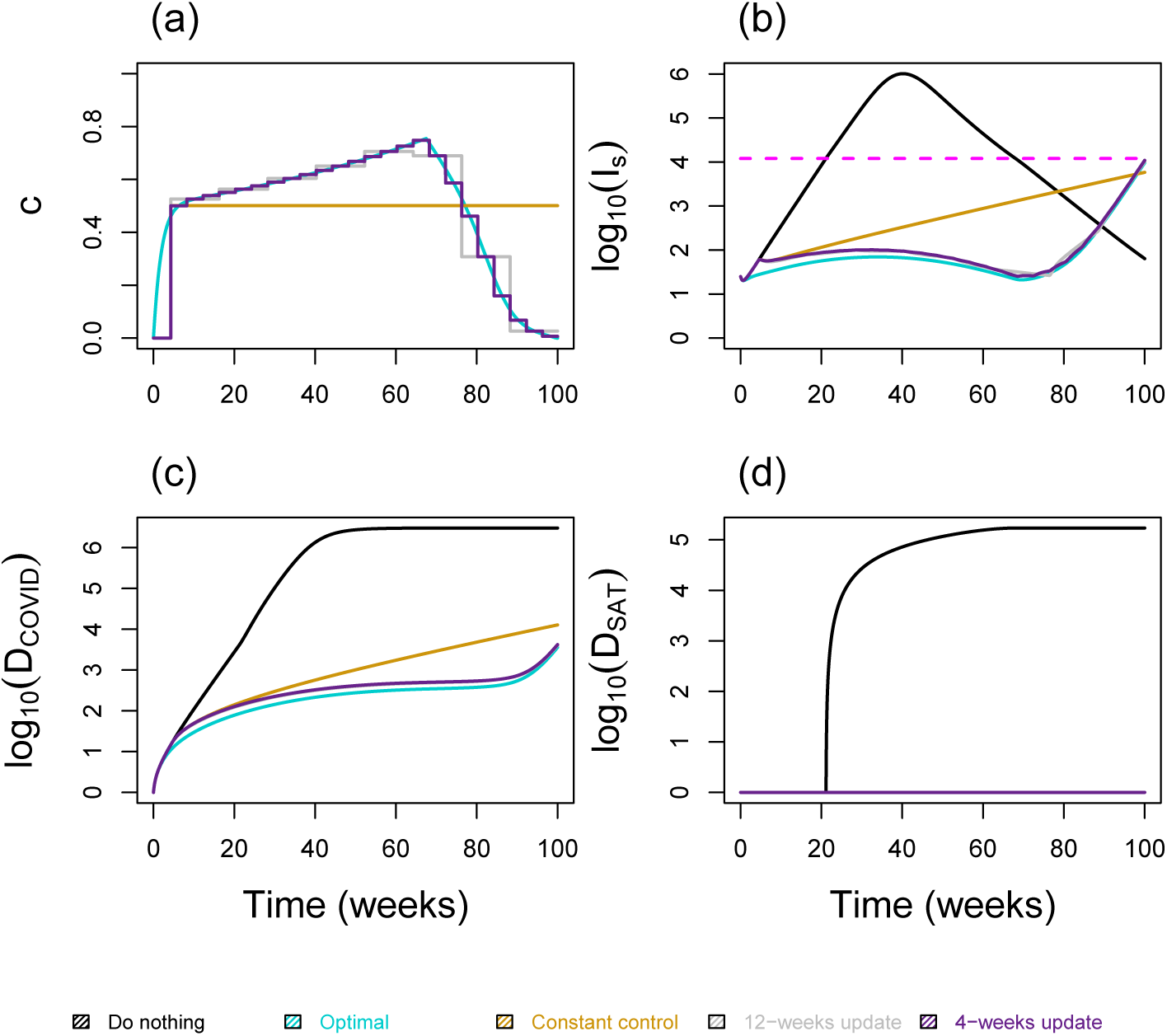
Effect of the optimal strategy and step constant control on the number of severe cases (*I_s_*), deaths directly attributable to COVID-19 (*D*_COVID_) and deaths indirectly linked to COVID-19 (*D*_SAT_). **(a)** Control strategies with *H* =1.2e+4 and *B* =4e+05. Step optimal controls are updated every 12-weeks (gray), 4-weeks (purple), or a unique period during which the mean of the optimal control is applied (gold). **(b)** Number of severe cases with and without controls. **(c)-(d)** Cumulative number of deaths with and without controls. Other parameters of the model are set to their reference values given in Table 1. The dash line represents the healthcare capacity (color online).

### Optimal control scenario

Overall, for our parameter values, the average of the optimal control during the control period is 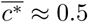. In the first stage, the optimal control increases rapidly to an intermediate value of approximately 0.5, followed by slow growth up to 0.65. (Figure 2a) and the epidemics remains under control for a long time period during which healthcare capacity is not overwhelmed (Figure 2b). Consequently, the number of deaths is strictly minimized during this first stage even though the control is far from its highest level (Figure 2c,d). After this stage during which a dampened epidemic peak is achieved, the number of severe cases starts to increase while remaining well below the healthcare capacity (Figure 2b), because the control intensity is progressively relaxed until the end of the time interval considered (Figure 2a). Importantly, in the absence of any take-over by health measures such as vaccine, the epidemic rebounces after the period of control. Indeed, only a small proportion of the population has been exposed (less than 0.3%) at the end of control period, which is far from the 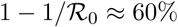 required to reach the herd immunity threshold.

Furthermore, the total number of deaths with this optimal scenario is substantially lower compared to the “doing nothing” scenario (Figure 2c,d). The cumulative number of deaths with the optimal control being at least 900 times smaller than when doing nothing. With the optimal control, the number of severe cases always remains below the healthcare capacity threshold, and thus the cumulative number of deaths indirectly attributable to the COVID-19 is minimized during the control period (Figure 2d). An “intuitive” rule that emerges from the optimal control (Figure 2a) is (i) from the beginning of the epidemic, gradually but rapidly increase control intensity to intermediate levels and then (ii) relax the control intensity gradually and slowly.

### The practicability of the optimal control

The optimal control is a continuous function and is then generally quite difficult to enforce in practice. Here, we show that based on the optimal control, we can derive step functions leading to potential practical implementations of the optimal control. We start with a “doing-nothing” period assumed to be a month. This period corresponds, for example, to the preparation time required for the implementation of a containment strategy. Next, the practical optimal control is updated every 4-weeks by keeping a constant amount of control during each 4-weeks period (Figure 2a, purple). The updating period can be relatively large, *e.g*., 12-weeks (Figure 2a, gray), or a unique period during which the mean of the continuous optimal control strategy is applied (Figure 2a, gold). Importantly, the constant defining the control intensity for each period is captured from the knowledge of the continuous optimal control strategy. With those step functions, the epidemic peak is strongly delayed compared to the scenario without any control (Figure 2b). The cumulative number of deaths at the end of the period of interest is strongly limited, both for the direct deaths caused by COVID-19 infections and for the indirect deaths caused by the saturation of the healthcare system (Figure 2c,d).

### Effect of the healthcare capacity *H*

By varying parameter *H*, we can illustrate the effect of healthcare capacities on the optimal control function. Small values of *H* correspond to situations where the control must be maintained for a very long time to preserve healthcare capacities (Figure 3a). As a consequence, corresponding optimal controls can be very intense to decrease severe cases and deaths (Figure 3b,c). On the other hand, large values of H correspond to situations where it is not necessary to maintain the health measure during all the control period. In this case, control intensities remain intermediate for relatively shorter period (Figure 3a), with similar infections and deaths compared to cases with smaller values of *H* (Figure 3b,c). However, we find that the shape of the optimal control function remains unchanged with an intense control followed by a decrease in control intensity.

**Figure 3:**
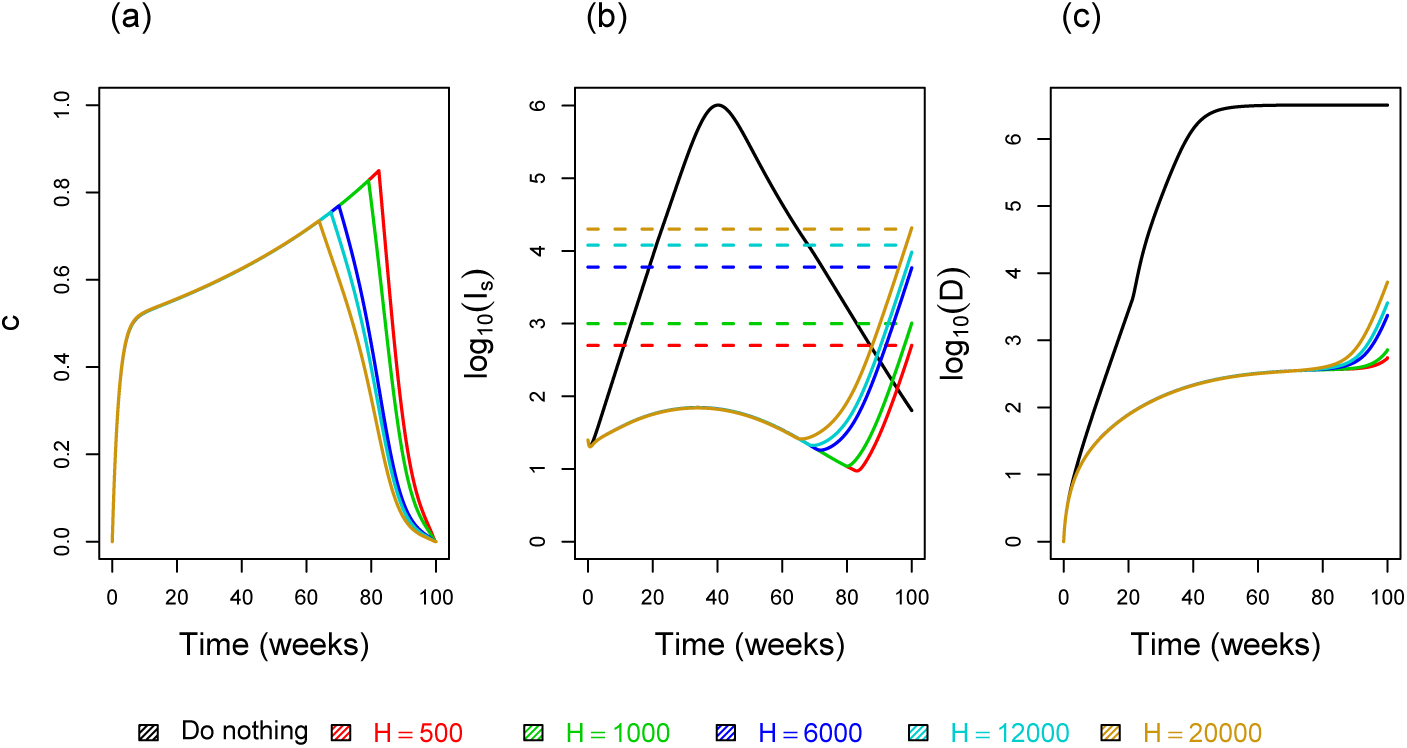
Effect of the healthcare capacity H on optimal interventions. **(a)** Control strategies. **(b)** Number of severe cases with and without controls. **(c)** Cumulative number of deaths with and without controls. Here we have *B ∈* {1.6; 2.16; 3.6; 4; 4.64} × (1e + 5) for *H ∈* {500;1000;6000;12000;20000} respectively, and other parameters of the model are set to their reference values given in Table 1. Dash lines represent the healthcare capacity (color online).

### Effect of the initial epidemic size on the optimal control

The initial size of the epidemic plays an important role in the optimal control strategy. The larger the initial number of infections, the higher the optimal control intensity (Figure 4a). Indeed, since the initial epidemic size is the number of cases at the time the decision is made to implement the containment strategy, the longer it takes to implement the strategy, the greater this size will be at the time the strategy is deployed. Consequently, the longer the delay in implementing non-pharmaceutical interventions, the tighter the control before the gradual decreasing. Also, the longer the delay in implementing the strategy, the less the optimal strategy can minimise the number of severe infections and deaths (Figure 4b,c).

**Figure 4:**
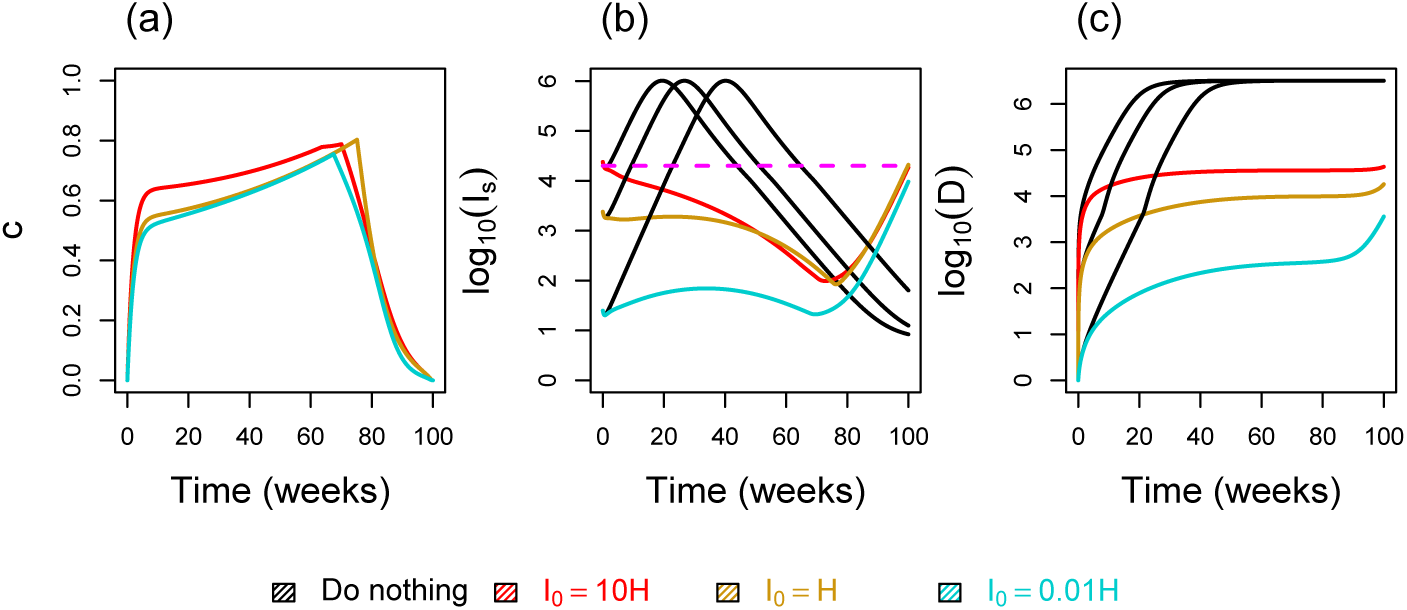
Effect of the the initial epidemic size *I*_0_ on optimal interventions. **(a)** Control strategies with *H* =1.2e+4 and *B* =4e+05. **(b)** Number of severe cases with and without controls. **(c)** Cumulative number of deaths with and without controls. Other parameters of the model are set to their reference values given in Table 1. The dash line represents the healthcare capacity (color online).

## 5 Discussion

The COVID-19 pandemics represents a dilemma for public health policies, which are faced with either mitigating the epidemic wave to rely on natural immunisation, or suppressing the wave long enough to develop and implement a vaccine. We focused on the latter case to address the following question: in a context where resources are finite, how to best allocate the control effort of the epidemic over the time period necessary to deploy a vaccine?

Using optimal control theory, we show that, assuming a quadratic cost for the control effort at a given time (*c*(*t*)), an optimal control strategy significantly reduces the number of deaths and is particularly sustainable at the population level. With this strategy, the intensity of the control increases rapidly to an intermediate value and then decreases steadily. Other strategies, such as implementing a strong short containment (“lock-down”) followed by a looser and constant control effort are almost comparable to a strategy without control in terms of cumulative number of deaths. Indeed, in this constant control strategy, the epidemic peak is delayed but barely dampened before the end of the control period.

As shown in the Appendix, we also investigated cycling strategies that alternate two levels (*c*_max_ and *c*_min_) of control intensity every 6 weeks. *c*_max_ is fixed to 90% for all scenarios and *c*_min_ is variable (Figure S1). In order to facilitate comparisons between scenarios, the area under the curve of each constant control is the same as for the optimal control *c*^*^. Overall, such scenarios delay the epidemic peak for the time during which controls are implemented. In the end of the period of interest, the cumulative number of deaths with cycling strategies remain generally more important compared to the optimal control. This configuration is the same with shorther cycles, *e.g*. a weekly cycle of 2 work days and 5 lock-down days (Figure S2).

Assuming an all-or-nothing scenario, we then considered an intense lock-down (*c* = 90%) with a variable duration of 12 or 50 weeks (Figure S3a). At first, this strategy maintains the epidemic dynamics under control (Figure S3b). However, as for the constant or the cycling strategies, these strategies only delay the epidemic peak and in the end the cumulative mortality over the time period of interest is comparable to the one without any control measure (Figure S3b,c).

These results should be interpreted in a qualitative sense because we have made a number of assumptions to derive our solutions. First, the structure of the model itself could be refined, for instance to explicitly model critical cases as in the model by [2]. Several epidemiological parameters of the COVID-19 epidemics also remain largely unknown at this stage [18]. For example, here we assume waiting rates to viral shedding (*ε*) and to symptom onset (*σ*) to be the same for severe and mild infections, which is not necessarily verified. The natural and disease-induced mortality functions could also be modified to fit even more to the data. For instance, both of these could depend on the intensity of the control *c* (*t*) because resource allocation to fighting COVID-19 can imply increased mortality by other diseases.

The question of controlling the epidemic before a deployment of a vaccine may also arise for other pharmaceutical interventions such as the development of a curative treatment. One of the limitations of our study is that we assumed that in the time necessary for vaccine development, the standard of treatment of COVID-19 infections remains constant. In reality, the time to discover and implement a new treatment could be lower than the time to discover and deploy a vaccine. This would not affect the general picture because the epidemic threat will remain unless the herd immunity threshold is reached. However, it would greatly affect the optimal strategy itself. Nevertheless, this same model remains valid in the context of treatment discovery if we redefine the time interval as the time necessary to set-up such a treatment. Qualitatively, we anticipate our results to hold on a shorter time interval. Indeed, assume that the control is to implement until a deployment of a pharmaceutical intervention within a year (approximately 50 weeks). In such configuration, we find that the optimal control leads to an increasing level of control with a maximum reached near the fourth month of the epidemics and a steady decrease until deployment of a such pharmaceutical intervention (Figure 5).

Our results offer new perspectives and research avenues to control the COVID-19 epidemics. In particular, we find that strategies that alleviate epidemic control too early all tend to only delay the epidemic wave. We also see that varying the level of control over time provides the best results for an overall given control effort over the period of interest. An open challenge is to identify optimal strategies that could be implemented in the field, especially by accounting for potential differences in the cost of control implementation among populations.

**Figure 5:**
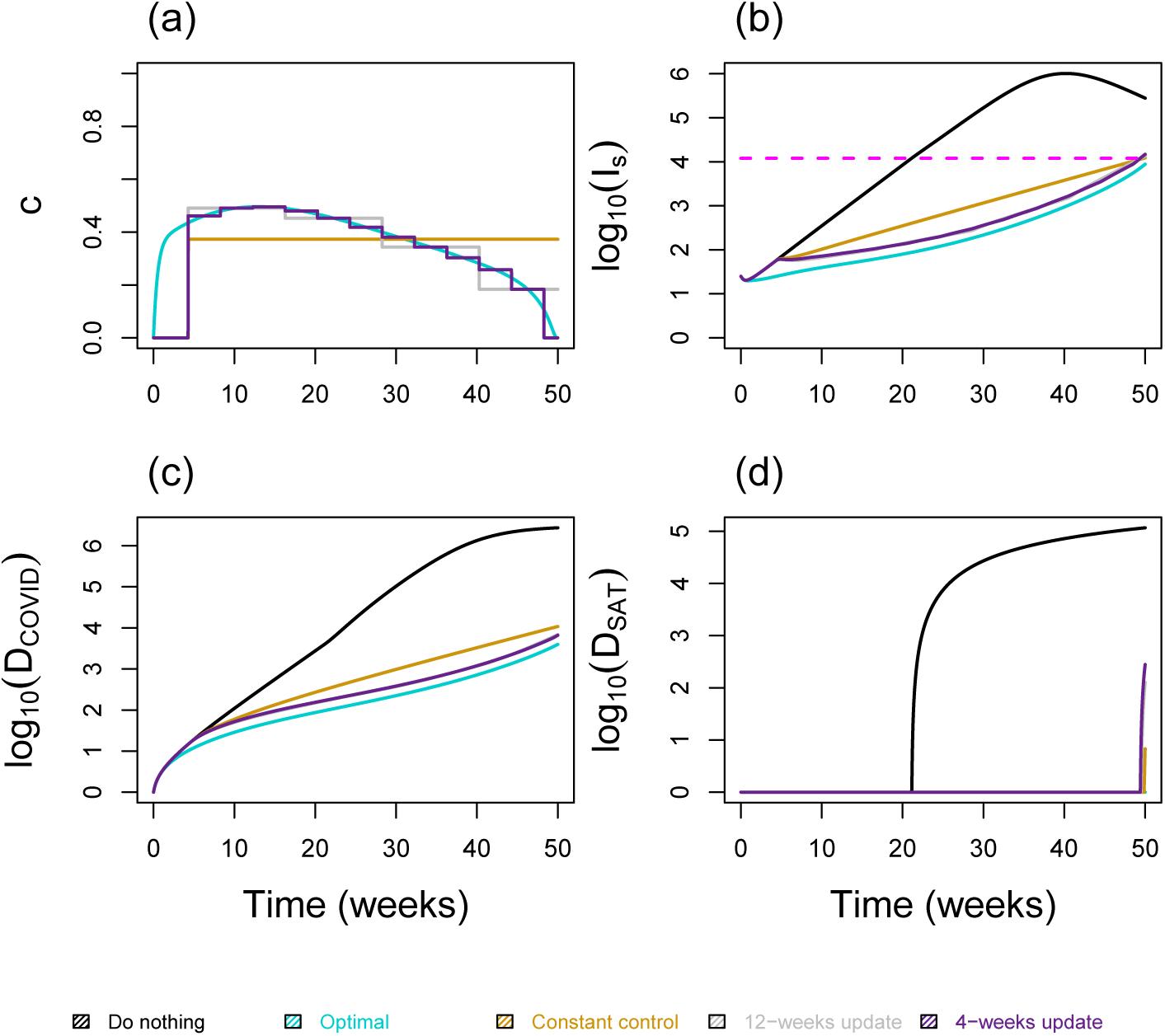
Optimal control during 50 weeks. **(a)** The control strategy with *H* =1.2e+4 and *B* =1.28e+05. **(b)** The number of severe cases. **(c)** The cumulative number of deaths. Parameters of the model are set to their reference values given in Table 1. The dash line represents the healthcare capacity (color online).

## Data Availability

All data pertaining to this study are available within the manuscript

## Acknowledgements

We would like to thank other members of the ETE modeling team: Thomas Bénéteau, Gonché Danesh, Baptiste Elie, Yannis Michalakis, Bastien Reyné, Quentin Richard and Christian Selinger.

## A Figures of cycling strategies that alternate two levels (*c*_max_ and *c*_min_)

**Figure S1:**
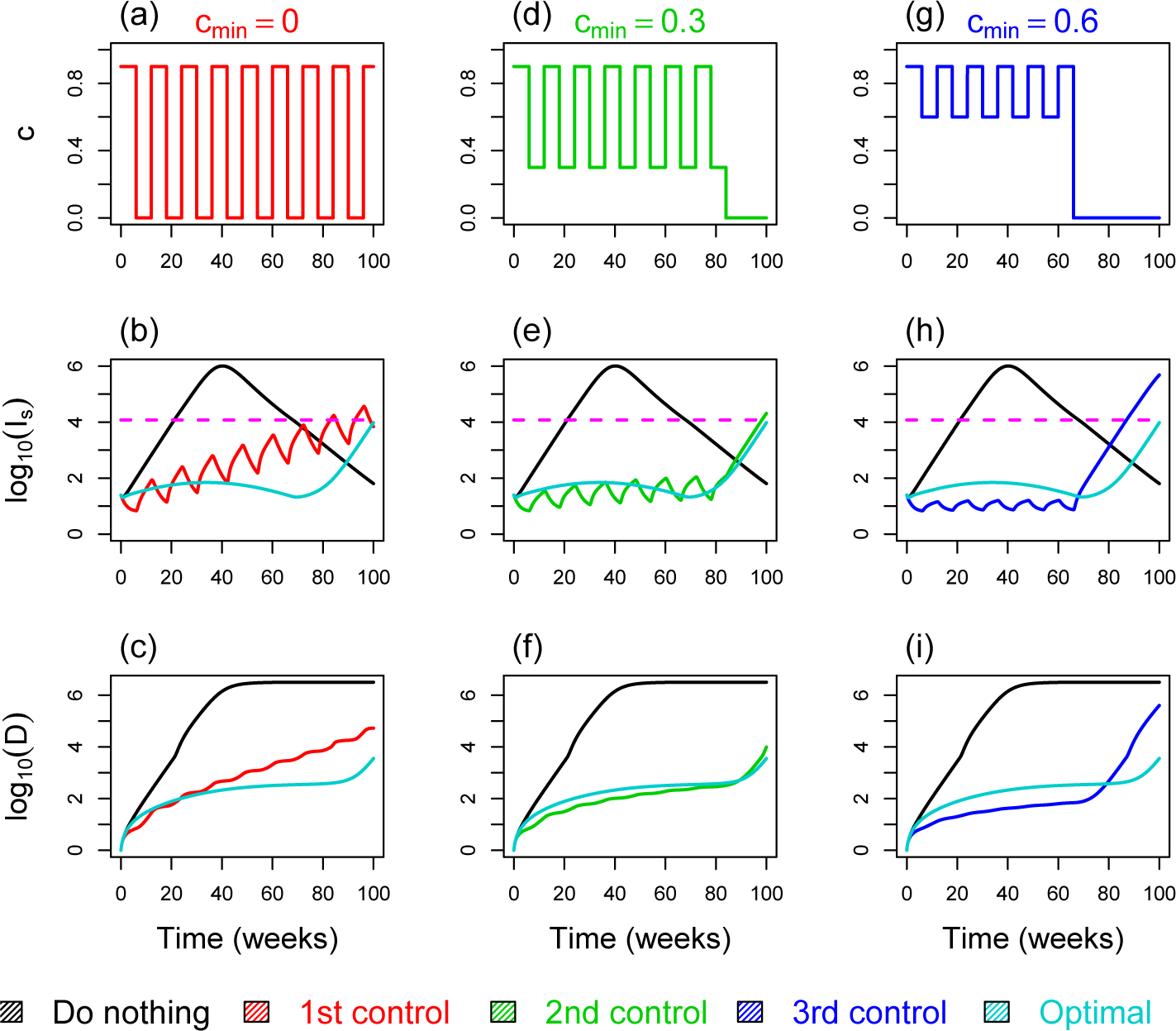
Intermediate constant control versus optimal control. In the intermediate control, each cycle last 12 weeks (with 6 weeks of lock-down at level *c*_max_ = 90% and 6 weeks at level *c*_min_). The first column **(a)-(b)-(c)** are respectively the control strategy, number of severe cases and cumulative number of deaths, with *c*_min_ = 0. Second column (d)-(e)-(f) with *c*_min_ = 0.3. Third column (g)-(h)-(i) with *c*_min_ = 0.6. Parameters of the model are set to their reference values given in Table 1. The dashed line represents the healthcare capacity (color online).

**Figure S2:**
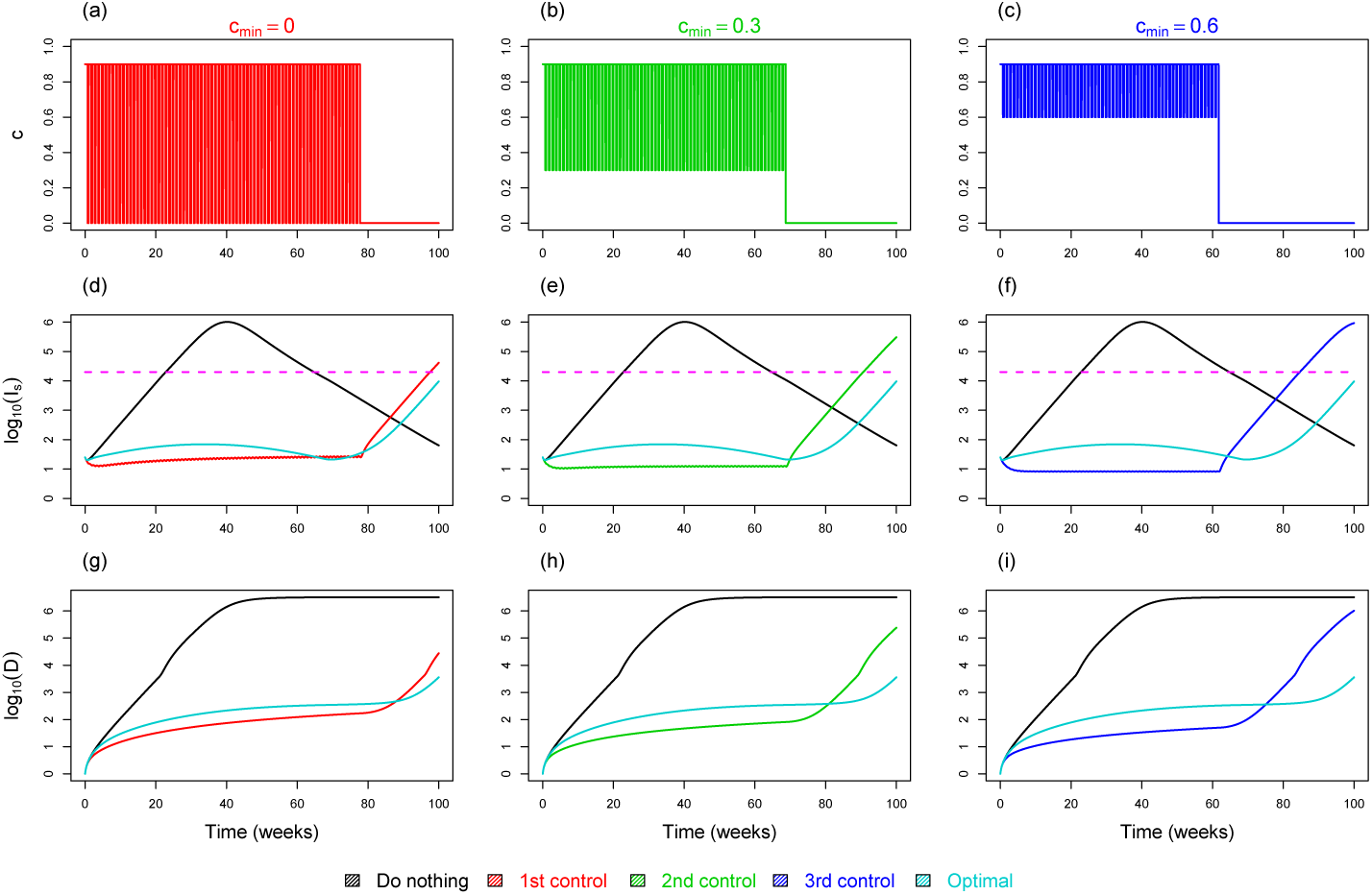
Intermediate constant control versus optimal control. In the intermediate control, each cycle last 1 week (with 5 days of lock-down at level *c*_max_ = 90% and 2 days at level *c*_min_). The first column **(a)-(b)-(c)** are respectively the control strategy, number of severe cases and cumulative number of deaths, with *c*_min_ = 0. Second column **(d)-(e)-(f)** with *c*_min_ = 0.3. Third column **(g)-(h)-(i)** with *c*_min_ = 0.6. Parameters of the model are set to their reference values given in Table 1. The dash line represents the healthcare capacity (color online).

## B Figures of full lock-down strategies

**Figure S3:**
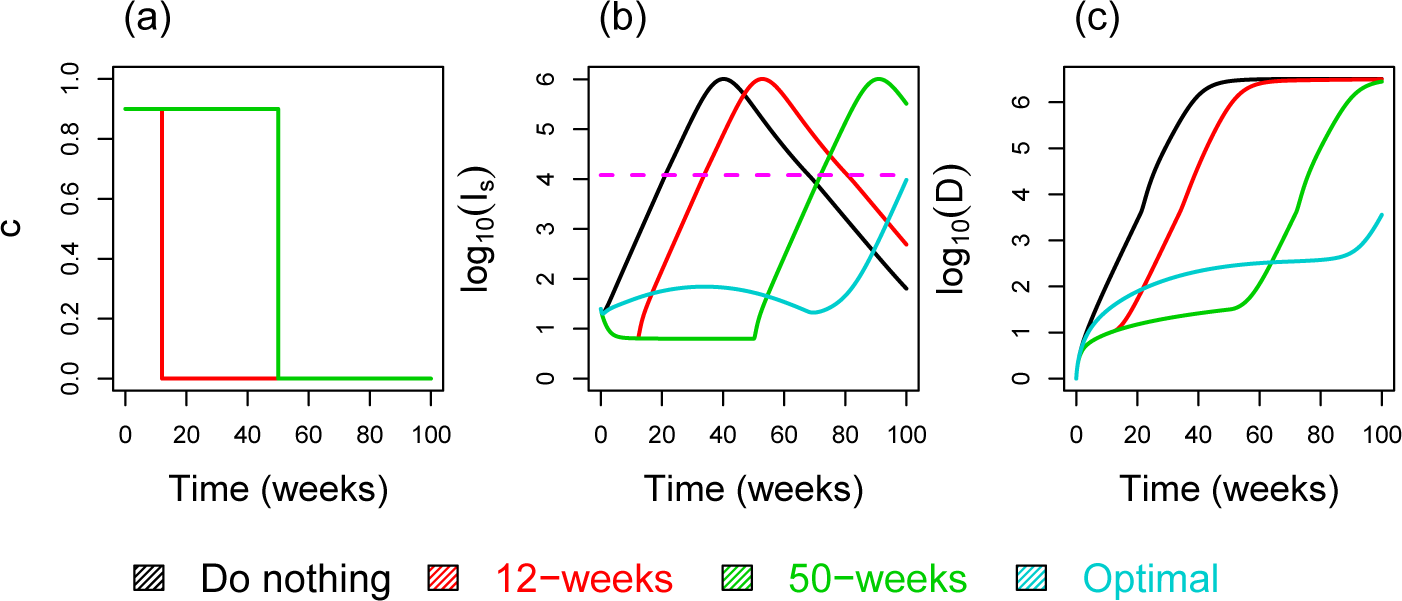
Full lock-down during 12 and 50 weeks versus optimal control. **(a)** The control strategy. **(b)** The number of severe cases. **(c)** The cumulative number of deaths. Parameters of the model are set to their reference values given in Table 1. The dash line represents the healthcare capacity (color online).

## C The necessary optimality condition

In order to deal with the necessary optimality conditions, we use Pontryagin’s maximum principle [22] and introduce the following Hamiltonian for the system (2.5)-(3.7)

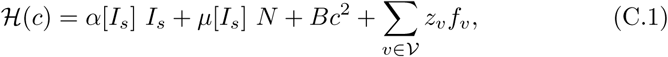

where 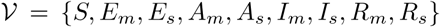; 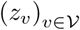 are adjoint functions and *f_ν_* is given by the right-hand side of (2.5) for the *ν*-compartment.

The necessary conditions for the existence of the solution to problem (3.7)

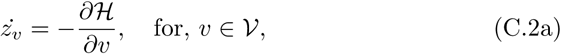

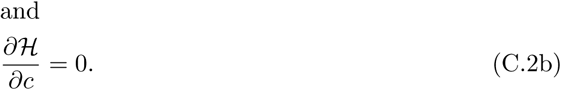

From (C.2a), we find that the adjoint system 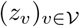 verify

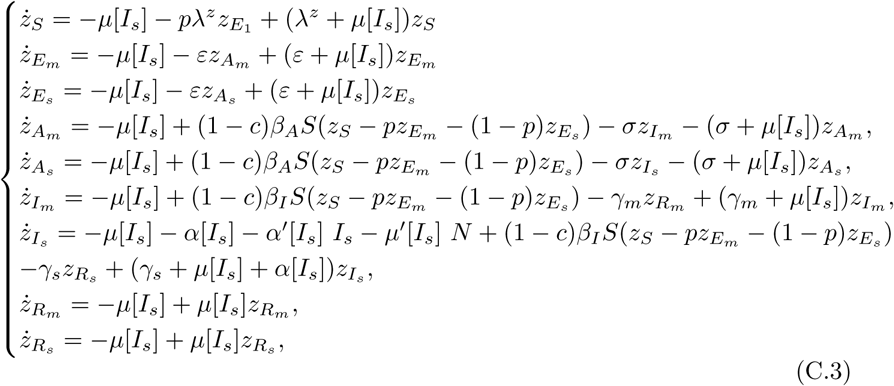

with the boundary conditions (or transversality conditions) *z_ν_* (*T*) = 0 and 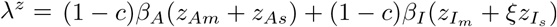.

If *c*^*^ is a solution of (3.7), then, following (C.2), it is characterized by

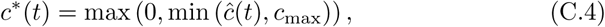

with

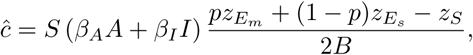

The proof of the existence of such control is standard and we refer to [8, 20] for existence results. Therefore, existence of the optimal control to the above problem is assumed.

## D The basic reproduction number

It is useful to write System (2.5) into a more compact form. To that end, we set *E* = (*E_m_*, *E_s_*), *A* = (*A_m_*, *A_s_*) and *I* = (*I_m_*, *I_s_*). As deaths and recovered are decoupled from the system, it is not necessary to keep them here. Here *x^T^* is set for the transpose of a vector or matrix x. Then, System (2.5) can be rewritten as

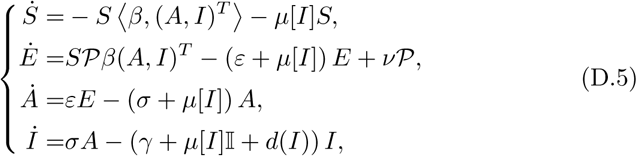

where *β* = (*β_A_*, *β_A_*, *β_I_*, *ξβ_I_*), 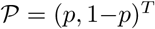, *γ* = diag (*γ_m_*, *γ_s_*), *d*(*I*) = diag (0, *α*[*I*]), 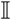 is the 2 × 2 identity matrix, diag (*w*) is a diagonal matrix which elements are given by w; and *x*, *y* is set for the usual scalar product of vectors *x* and *y*.

Linearizing System (D.5) at the disease free environment (*S, E, A, I*) = (*S*_0_, 0,0,0) and setting *u* = (*E*, *A*, *I*)*^T^*, we obtain 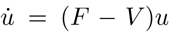, where *F* is the rate of appearance of new infections in each class, and *V* is composed by the rate of transfer into each class by all other means and the rate of transfer out of each class. It the comes

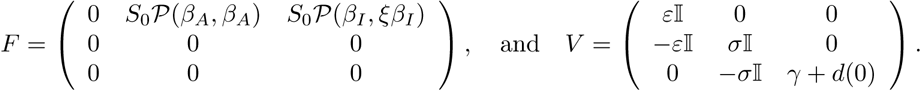

Following [4], the basic reproduction number of Model (2.5) is defined as the spectral radius *ρ* (*FV*^−1^) of the next generation matrix *FV*^−1^. A straightforward computation leads to

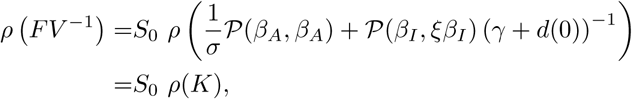

with 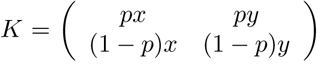, 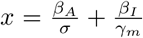 and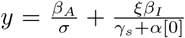. Since det(*K*) = 0, it comes

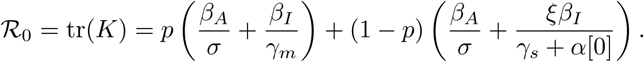

